# A study on RSR evaluation of health service capacity of county-level administrative areas based on entropy weight TOPSIS method in Fujian Province in China

**DOI:** 10.1101/2022.10.11.22280944

**Authors:** Shengli Zhang

## Abstract

**Objective:** To identify the differences in health service capacity among county-level administrative regions in Fujian province in China, so as to provide decision-making ideas and basis for high-quality development of the county medical community system with Fujian characteristics,and to prevent health reform from diverging from the essence of meeting everyone’s health needs and improving health service capacity.

**Methods:** The entropy weight TOPSIS and RSR methods were used to comprehensively evaluate the health service capacity of 84 county-level administrative areas in Fujian Province in China.All data are from the official website of Fujian Provincial Bureau of Statistics.

**Results:** The results show that the evaluation system of health service capacity with seven indicators, including per capita GDP, number of resident population, number of health institutions, number of beds in health institutions, number of health technicians, number of licensed (assistant) doctors and number of registered nurses,has a good discrimination. The index entropy weights are 0.09, 0.11, 0.12, 0.15, 0.17, 0.19, 0.17 respectively.According to the principle of reasonable grading of normal distribution, the health service capacity is divided into three grades by the Probit value, including 13 weak ones and 14 strong ones. It is worth noting that, as a typical example of China’s health reform, the county-level health service capacity of Sanming City in Fujian Province is only in the middle, and even Ninghua County is weak.

**Conclusions:** The entropy weight TOPSIS and RSR methods make use of the characteristics of the data itself, avoid the influence of subjective judgment of the evaluation subject, and can quickly make classification judgment. This study uses the statistical yearbook data to construct an evaluation model of 7 indicators, which can well classify the health service capacity of 84 county-level administrative areas. According to the evaluation results of the health service capacity in Sanming City, where the health reform model has been widely praised, it suggests that the health service capacity building cannot be ignored in the health reform evaluation. An objective evaluation may provide more valuable reform recommendations.

## 1. Introduction

Health service capacity is one of the primary conditions to promote the development of health cause.Any country or region’s macro health decision-making can not be separated from scientific research and accurate judgment of health service capacity. The health reform of Fujian Province has always been at the forefront of China’s health reform, among which the Sanming medical reform model has been affirmed by the central leadership and strongly recommended by the government departments. To promote the high-quality development of health services and deepen the quality of Fujian medical reform model, it is necessary to re-examine the health service capacity of each county-level area in Fujian province at the present stage. The low capacity of health services has a major impact on the development of health policies and the effectiveness of health reforms. But the comprehensive evaluation index system of health service capacity has not unified common understanding. There are many algorithms to calculate the index weight, and there is always controversy between objective and subjective weight determination.Therefore, in this study, a health service capacity evaluation index system with consensus indicators was constructed, and the data of these indicators could be easily obtained from the official website. According to the index system, the objective entropy weight method, TOPSIS method and RSR method were combined to grade and classify the health service capacity of 84 county-level administrative areas in Fujian Province in China,and some suggestions for health service capacity evaluation and decision-making were put forward.

## 2. Data and methods

### 2.1 Data source

All data in this study are from “Fujian Statistical Yearbook 2021” published on the official website of Fujian Provincial Bureau of Statistics.The data is public and open. The data related to health service capacity of 84 county-level administrative areas in Fujian Province were collected and analyzed.

### 2.2 Construction of evaluation index system

The evaluation of health service capacity belongs to the resource-based evaluation. As a resource-based evaluation, its research scope includes the evaluation of human resources, financial resources, material resources, transaction handling capacity, informatization level, technology level, etc. According to literature research, most researchers [1-4] used consensus indicators to analyze health service capacity by using yearbook data, mainly including the number of hospital institutions, the number of beds, and the number of health technicians per unit population. Some scholars [2-4] also included indicators such as the number of people diagnosed and treated, diagnosis and treatment expenses, hospitalization expenses, and bed utilization rate into the analysis. Because the number of patients, cost and bed use are outcome indicators of health services and belong to the scope of health service utilization evaluation, the evaluation indicators adopted in this study, after expert consultation, finally determined the indicators include the number of health institutions, the number of beds in health institutions, the number of health technicians, licensed (assistant) physicians, registered nurses and other consensus indicators. In this study, the per capita GDP index is specially introduced to reflect the regional economic level. It is generally believed that the regional economic level is positively related to the cost invested in the health field. The indicator of permanent population is also introduced. It is generally believed that the population is negatively related to the capacity of health services. With other resources unchanged, the effective supply capacity of health services will decline when the population increases.

### 2.3 Research methods

In this study, entropy weight TOPSIS and RSR method were used to comprehensively evaluate the health service capacity of 84 county-level administrative areas in Fujian Province, and some suggestions for health service capacity evaluation and decision-making were put forward.Excel software was used to compile a modular process to calculate entropy weight and Ci value of TOPSIS model, and IBM SPSS25 was used to verify RSR distribution, calculate regression equation and correlation coefficient. Finally, Excel function was used to rank and classify the evaluation result data.

#### 2.3.1 Entropy weight method

Entropy weight method [5-10] is a quantitative weighting method that does not rely on expert opinions, but mining the characteristics of data itself, with objective characteristics. Based on the variation degree of the index value, the entropy weight is calculated by using the information entropy to correct the index value. The greater the degree of variation, the more information contained, the smaller the entropy, and the greater the weight. On the contrary, the weight is smaller.If the value of an indicator is equal, the indicator has no variation and the weight is zero. The importance of each decision-making indicator can be reflected by the difference of information contained in each indicator value. The analysis process can be realized with EXCEL software.

Entropy weight follows the procedure below. The original data matrix 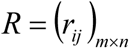 is formed by *n* indicators of *m* things to be evaluated, where *r*_*ij*_ is the jth indicator value (*j*=1, 2,…, *n*) of the ith thing to be evaluated (*i*=1, 2,…, *m*). First of all, the index values of the things to be evaluated are treated in the same direction or trend. Positive indicators (such as benefit type and output type) are calculated according to Formula 1, while negative indicators (such as cost type and input type) are calculated according to Formula 2. Secondly, the index value after the same direction is standardized according to Formula 3. Thirdly, calculate the entropy value of the jth index according to Formula 4. Finally, calculate the entropy weight of the jth index according to Formula 5.Formulas 1 to 5 are as follows.

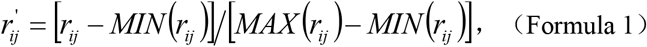

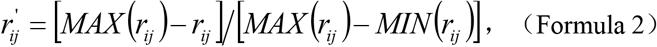

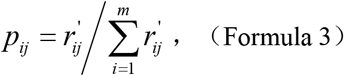

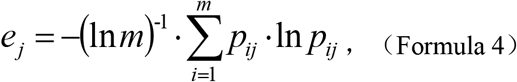

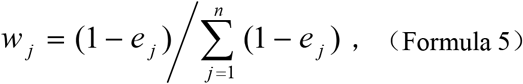

Obviously, the entropy weight is affected by the number of indicators and the number of evaluation objects. Therefore, the entropy weight method is applicable to the index weighting when evaluating a given number of objects, which belongs to variable weight and is not affected by the will of the evaluation subject.

#### 2.3.2 TOPSIS method

TOPSIS (Technique for Order Preference by Similarity to Ideal Solution) is an evaluation method for limited schemes of multi index decision-making. It uses the information of original data to reflect the gap between evaluation schemes. There is no requirement on data distribution type and sample size [11]. Based on the normalized matrix, the optimal scheme and the worst scheme in the finite scheme are found out, and the distance between the index value of each thing to be evaluated and the optimal scheme and the worst scheme is calculated. Finally, the relative proximity between the things to be evaluated and the optimal scheme is calculated as the basis for the good.

TOPSIS evaluation follows the steps below. First of all, the actual measurement or actual evaluation value of the original data matrix 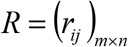 formed by *n* indicators of *m* things is processed in the same direction. Generally, negative indicators (such as cost type and input type) are counted backwards. Secondly, the index values after homoorientation are normalized according to Formula 6 to form a new normalized matrix 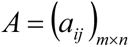 and to realize dimensionless. Thirdly, according to the normalized matrix, the optimal value vector 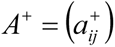 and the worst value vector 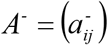 are obtained, where 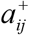 represents the maximum value of the jth index of the *m* things to be evaluated, and 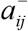 represents the minimum value. Fourthly, the distance between the *m* things to be evaluated and the optimal vector is calculated according to Formula 7, and the distance between them and the worst vector is calculated according to Formula 8. Finally, the proximity of *m* things to be evaluated and the optimal vector is calculated according to Formula 9. The *m* things to be evaluated are sorted according to their proximity, and the larger the value, the better the evaluation.Formulas 6 to 9 are as follows.

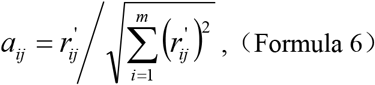

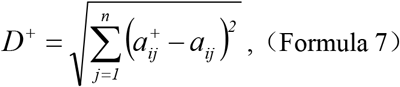

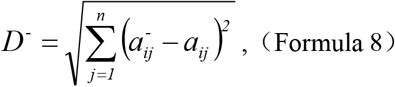

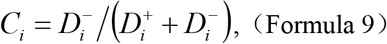

#### 2.3.3 RSR method

RSR(Rank Sum Ratio) refers to the average number of row (or column) ranks, which is a nonparametric statistic with (0, 1) continuous variable characteristics. The RSR synthesizes the information of multiple evaluation indicators, indicating that the higher the RSR, the better the comprehensive level of multiple evaluation indicators. RSR evaluation follows the steps below. Firstly, the proximity between the health service capacity of 84 county-level administrative areas and the optimal vector was taken as the RSR value, and the frequency, cumulative frequency, average rank and cumulative frequency of different groups were calculated from the smallest rank to the largest rank. The last cumulative frequency was calculated by “(1-1/4*m*)×100%”. Secondly, the cumulative frequency is taken as the independent variable, and the EXCEL expression “5+NORMSINV()” is used to calculate the probability Probit. Thirdly, the correlation coefficient and regression equation were calculated with Probit as the independent variable and RSR value as the dependent variable. According to the principle of normal distribution and reasonable classification, the Ci of 84 county-level administrative areas were divided into three grades according to the estimated value of RSR corresponding to Probit values 4 and 6.

## 3. Results

### 3.1 Comprehensive evaluation results of entropy weight TOPSIS model

The entropy weigh of 7 health service capacity indicators of 84 county-level administrative areas calculated by TOPSIS comprehensive evaluation method is 0.09, 0.11, 0.12, 0.15, 0.17, 0.19 and 0.17 respectively.The order is made according to the *C*_*i*_ value, and the specific ranking is shown in Table 1.

**Table 1.**
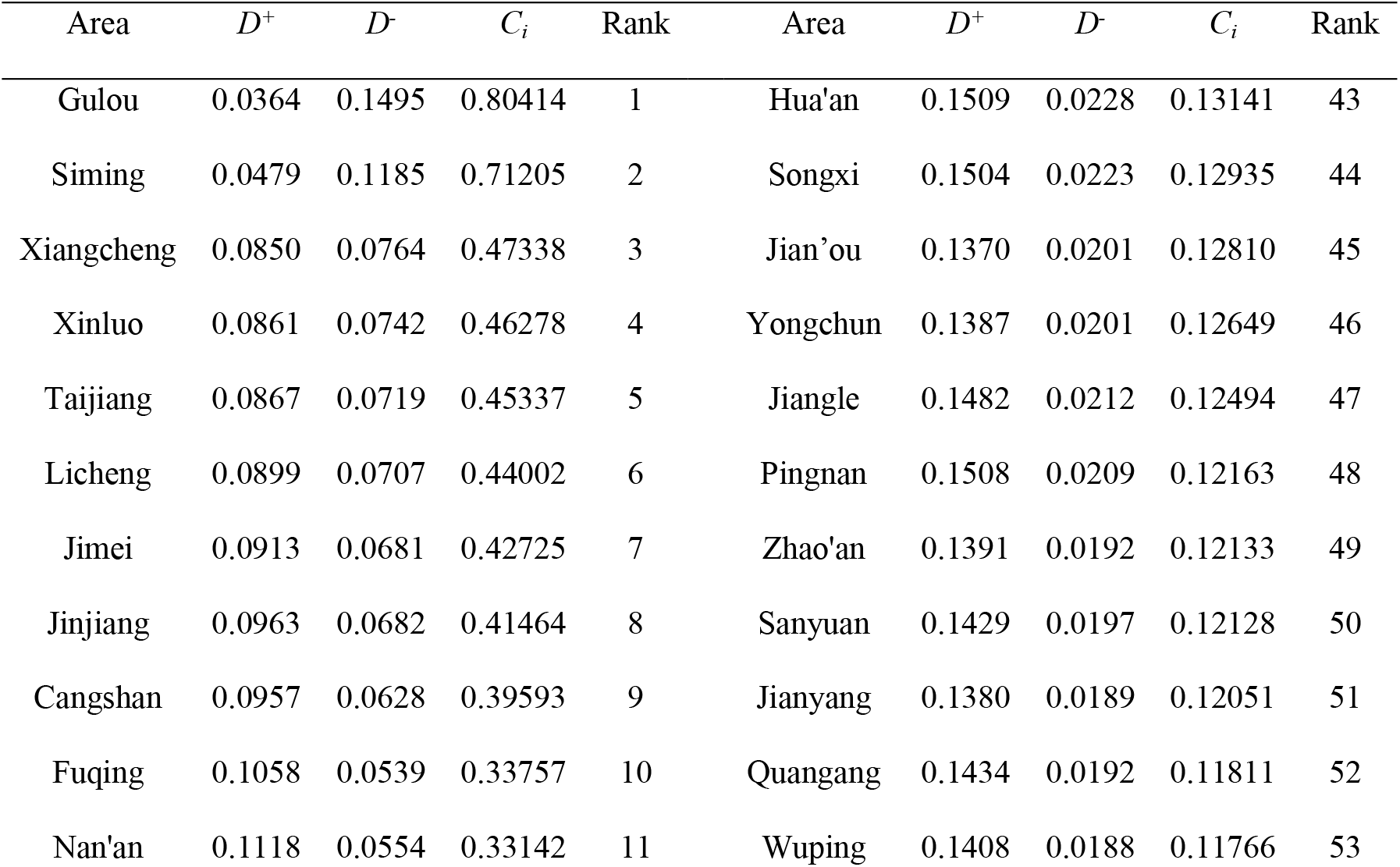
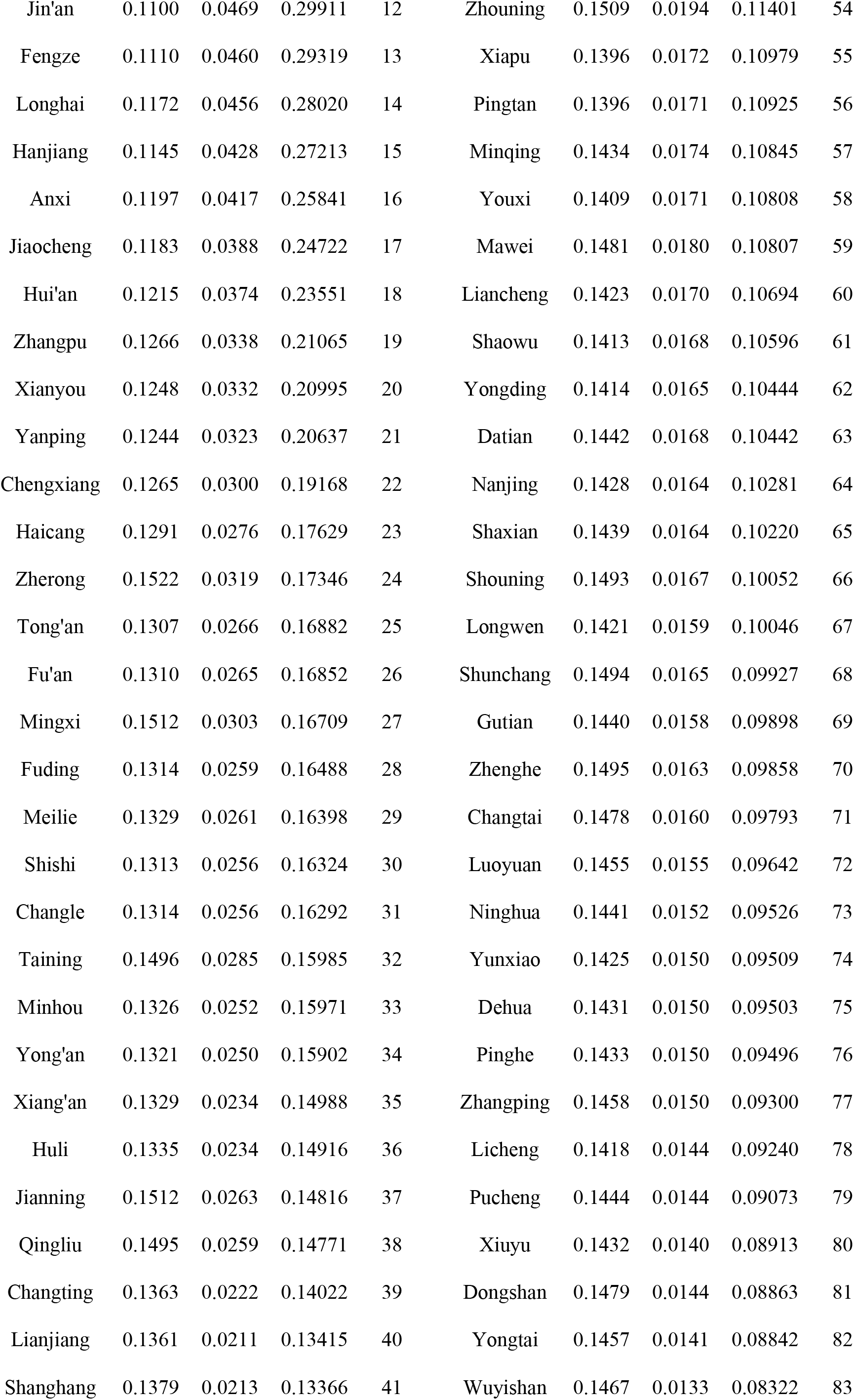
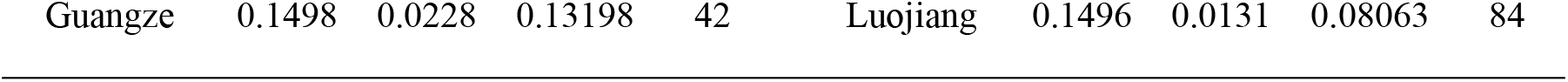
Comprehensive evaluation results and ranking of entropy weight TOPSIS model.

### 3.2 Probit evaluation results of RSR model

The RSR value of health service capacity in 84 county-level administrative areas is expressed by its *C*_*i*_ value, and the Probit value is shown in Table 2. The *C*_*i*_ value of the health service capacity in 84 county-level administrative areas was linearly related to the Probit value (*R*^2^=0.717, *P*<0.001), and the regression equation *C*_*i*_=0.114 *Probit*-0.39 (*F*=207.932, *P*<0.001). According to the Probit value of the health service capacity in 84 county-level administrative areas, the health service capacity in 84 county-level administrative areas is divided into three grades, including weak (<4), general (4–6), and strong (>6) according to the principle of reasonable classification of normal distribution. Among them, the weak include 13 county-level administrative areas, and the strong include 14 areas. The specific classification is shown in Table 3.

**Table 2.**
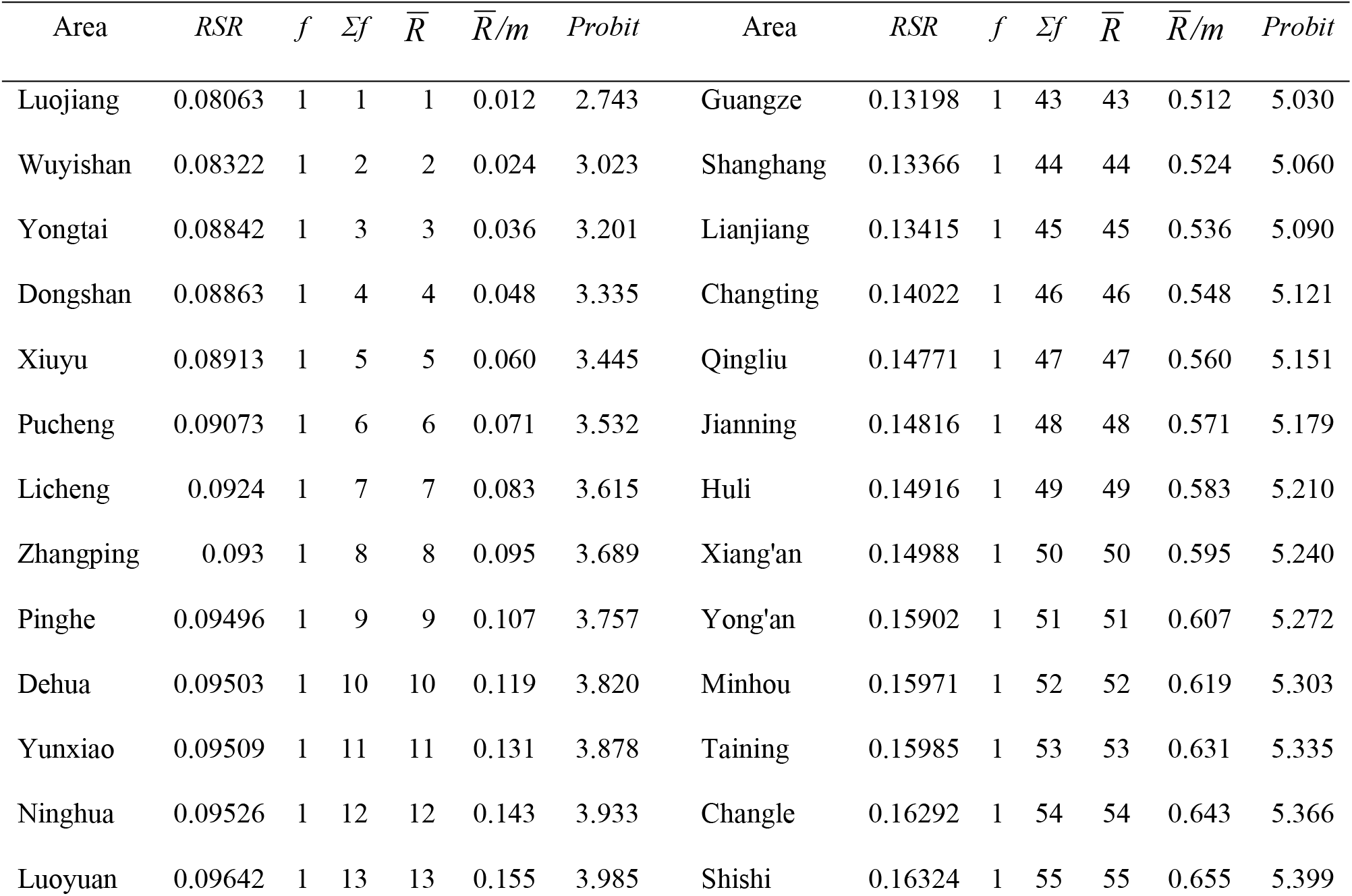
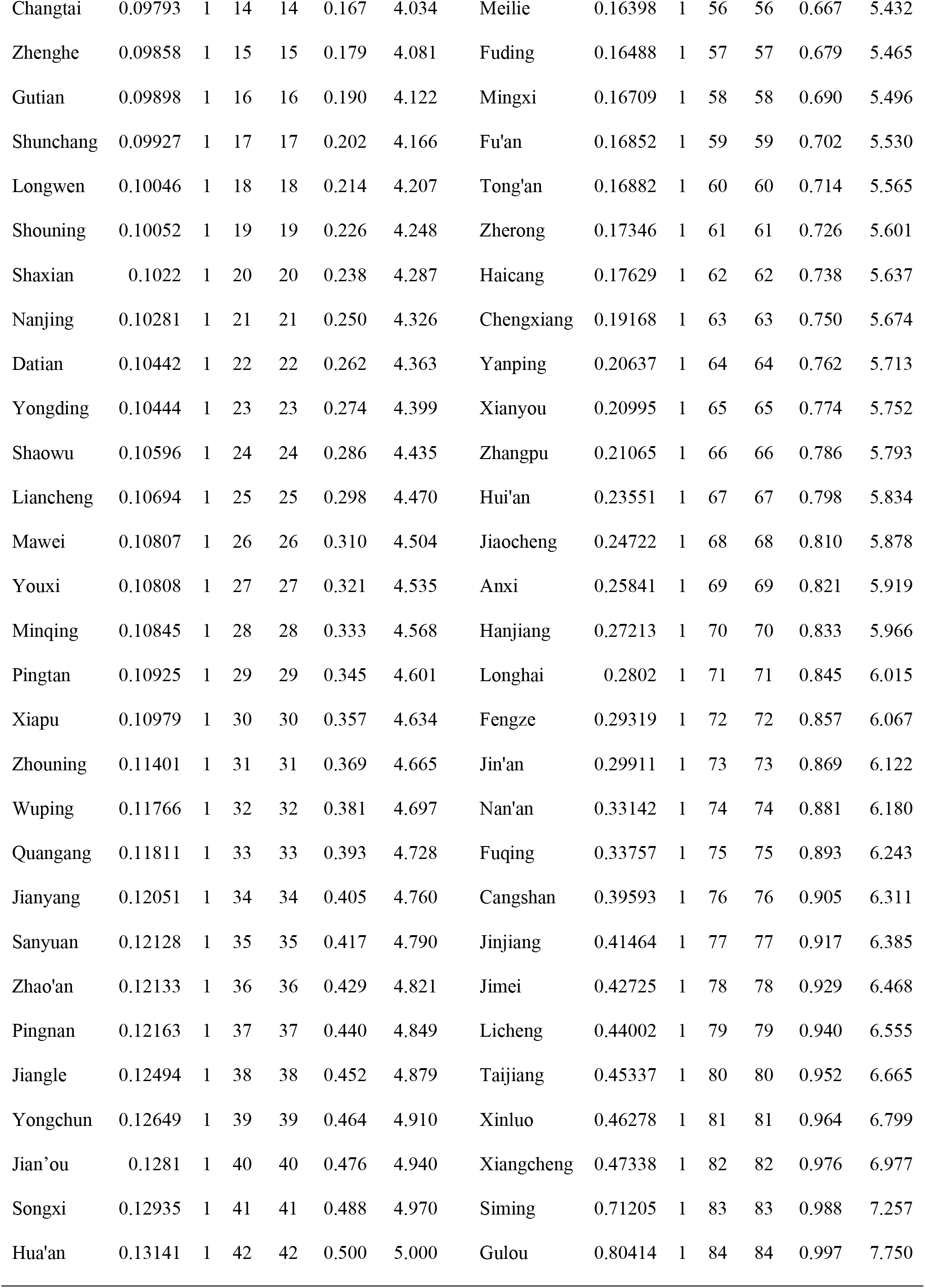
Probit evaluation results of RSR model of health service capacity in 84 county-level administrative areas.

**Table 3.**
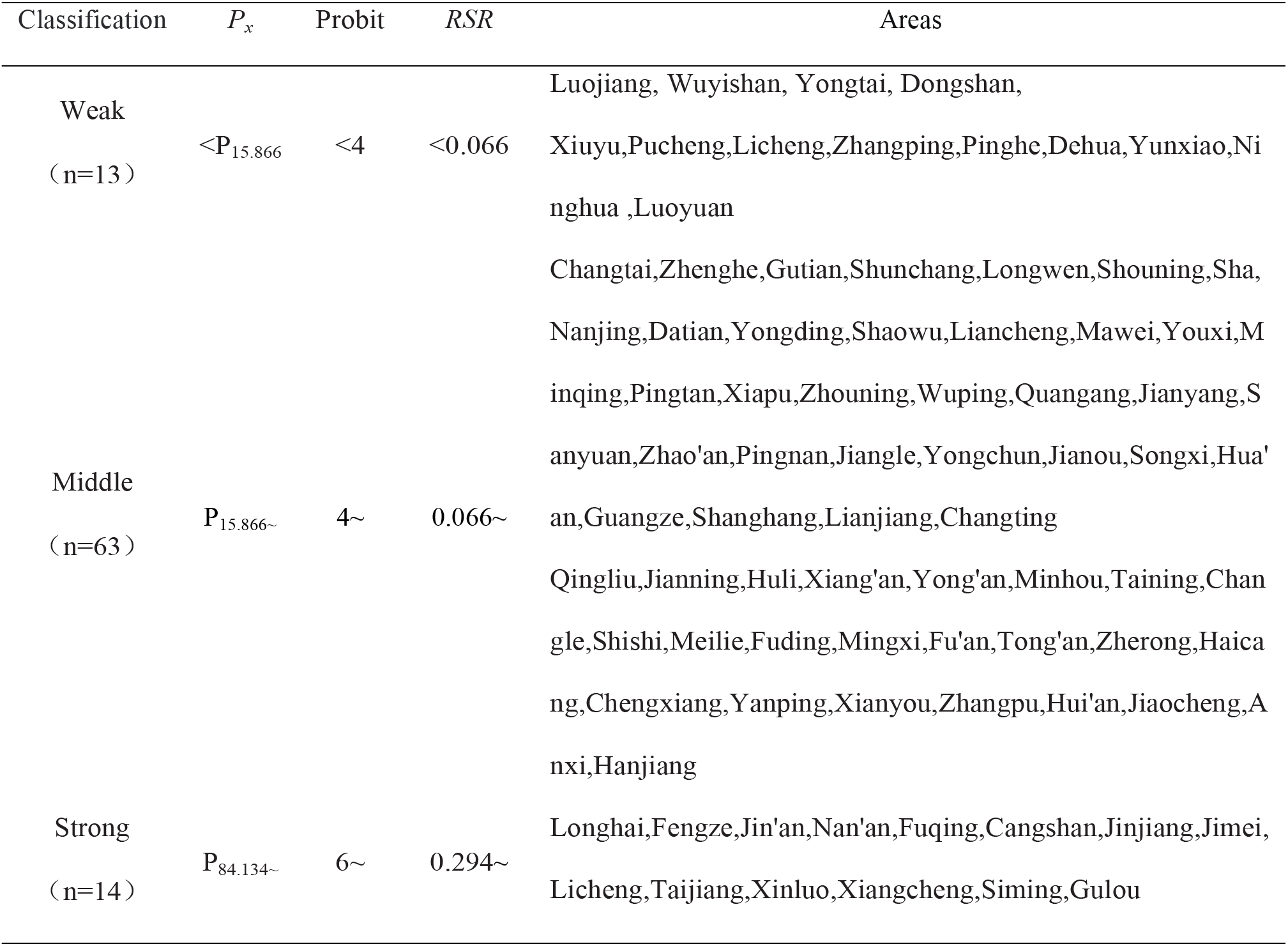
Three-level classification of health service capacity in 84 county-level administrative areas.

## 4. Discussions

The seven index evaluation system of health service capacity has a good ranking role. It can be seen from the ranking results of entropy weight TOPSIS that the evaluation system of health service capacity, which includes seven indicators: per capita GDP, number of resident population, number of health institutions, number of beds in health institutions, number of health technicians, number of licensed (assistant) physicians, and number of registered nurses, has a good differentiation. This indicator set directly uses the original data for calculation and modeling, without the calculation of relative numbers, so the data information has high fidelity. According to the ranking of this indicator set, 13 of the 84 county-level administrative areas have weak health service capacity, put forward higher demand for financial input, and higher demand for human resources than other areas. Therefore, corresponding policies and measures should be taken to improve the health service capacity in these areas. It is particularly pointed out that the health service capacity of 12 county-level administrative areas of Sanming City, including Ninghua County, Shaxian County, Datian County, Youxi County, Sanyuan District, Jiangle County, Qingliu County, Jianning County, Yong’an City, Taining County, Meilie District and Mingxi County, is evaluated as “weak” or “average”.This evaluation result is different from the previous positive evaluation of Sanming medical reform. This shows that, at least in terms of health service capacity, the quality improvement of the medical reform needs to be strengthened.

RSR modle ranking is based on the regression relationship of the measured values to carry out equation fitting, and the sample information is used to estimate the overall characteristics. Combining the characteristics of probability unit value to rank according to the predicted value is more reasonable than directly using the measured value, which makes the decision guidance value better.RSR ranking can better indicate the priority strategy of health policy.

Through the analysis, the results show that there is a significant gap in the health service capacity of 84 county-level administrative areas. This gap is not only reflected in the differences brought by GDP, but also in the “coastal” and “inland” gaps. It is not only reflected in the number of people served, but also in health human resources. All of these suggest that health fiscal policies cannot be determined simply by economic level, nor can they be treated simply by “coastal” or “inland”. It is also not allowed to take the municipal districts as the basis for macro policies. Scientific decision-making needs scientific evidence, and comprehensive evaluation of health service capacity is an important scientific basis for health policy.The high-quality development of the County Medical Community with Fujian characteristics, from “controlling medical expenses” to “taking health as the center”, must be based on scientific decision-making, and it is an insurmountable step to scientifically identify the differences in health service capacity of County Medical Community. It is strongly recommended that the decision-making department further excavate health big data resources, comprehensively study and judge from multiple perspectives and fields, such as health service capabilities, so that digital can truly enable the development of health undertakings.All evaluations and decisions should be based on objective and scientific evidence as far as possible, at least including objective and scientific data sources and analysis methods, rather than personal likes and dislikes.It is specifically stated that this study does not exclude the importance of scientifically designed qualitative research based on expert consultation, such as the Delphi method and the thematic meeting method, and does not deny the value of the weight coefficient of the analytic hierarchy process.

## Data Availability

http://tjj.fujian.gov.cn/tongjinianjian/dz2021/index.htm

## Acknowledgement

The study was supported by grants from Major Project sponsored by Fujian New Think Tank (21MZKB10), Fujian Provincial Department of Education Project(JAS20113), and Fujian University of Traditional Chinese Medicine First-class Undergraduate Course Project (Minzhong Medical Education [2021] No. 46)

